# Evidence of rainfall-driven disruptions in respiratory epidemics surveillance

**DOI:** 10.1101/2025.08.05.25333040

**Authors:** Susanna Caruso, Claudio Ascione, Eugenio Valdano

## Abstract

Extreme weather events can disrupt human behavior and access to health services, possibly affecting the surveillance of respiratory epidemics. Rainfall, in particular, can reduce the uptake of diagnostic testing, delay the detection of infections, and amplify the disadvantages faced by vulnerable populations. We examined this effect using the COVID-19 pandemic as a case study, analyzing daily testing activity in mainland France and New York State between May 15, 2020, and November 1, 2021. COVID-19 testing data were combined with daily rainfall and local socioeconomic indicators. Epidemic trends and weekly seasonality were extracted using time series modeling and incorporated into Poisson regression models to quantify the impact of rainfall on testing rates and to explore its variation with local rainfall profiles and income levels. Rainfall was consistently associated with reduced testing in both France and New York State. Nationally, 25 millimeters of rainfall in one day were linked to an 8-to-9 percent reduction in testing in France and around a 5 percent reduction in New York State. Spatial patterns revealed that decreases were more pronounced in poorer areas, while locations accustomed to frequent light rainfall showed weaker effects. These findings, across diverse geographies and climates, show that rainfall can hinder respiratory epidemic surveillance by reducing testing, and compounds socioeconomic and climatic vulnerabilities. Recognizing and mitigating weather-driven disruptions is essential to strengthen epidemic surveillance and maintain timely detection under a changing climate.

## Introduction

Climate change is driving a rise in both the frequency and intensity of extreme weather events^1^. These events impact human health through multiple pathways^2^, and notably by altering the transmission dynamics of respiratory pathogens. One driver of this is the disruption of human behavior and social mixing patterns during and after extreme events^3^. Studies showed that severe snowfall and rainfall were associated with a reduction in community contact rates and, through that, COVID-19 incidence^4,5^, while heatwaves and wildfire had the opposite effect by increasing the amount of time spent indoors and thus the likelihood of airborne transmission^6,7^. Wildfires and COVID-19 were also proven to have a compound effect on excess mortality^8^. Also, severe floods were associated with a wider spatial spread of emergent respiratory pathogens^9^. More generally, the relationship between extreme weather and epidemic circulation is complex, reflecting the ways in which such events disrupt human mobility and contact patterns. Weather affects daily routines, reshapes social interactions^10–13^ and alters the way individuals engage with points of interest, such as workplaces, recreational areas, healthcare facilities or shelters^3,7^. Crucially, these behavioral shifts can influence pathogen transmission both directly, by modifying contact networks, and indirectly, by undermining the public health response. Disruptions to service access^14^ and infrastructure, or risk perception, may delay the detection of cases, hamper access to healthcare^15^ and hinder the implementation of preventive and containment measures. When extreme weather coincides with epidemic emergencies, it can thus compromise the effectiveness of public health campaigns, on top of its direct effect on circulation^2^. And as climate change increases the likelihood of such compound events, it becomes essential to understand their impact to inform preparedness and response strategies.

In this study, we examined whether heavy rainfall during the early COVID-19 epidemic was associated with reductions in testing rates in the state of New York and in France. We quantified the strength of this association and assessed its geographic variability. The COVID-19 pandemic generated an unprecedented volume of data, offering detailed insight into transmission dynamics, testing behavior, and public compliance with health measures^16–18^. These datasets have proven critical for evaluating the effectiveness of interventions^19–21^. A retrospective analysis of how rainfall affected the public health response during this large-scale respiratory epidemic will thus contribute to the broader effort of preparing for future outbreaks of emerging or re-emerging pathogens under a changing climate.

## Methods

### Data

We selected Admin-2 as the geographic resolution of our analysis, corresponding to departments (France) and counties (New York State). In France, we included the 94 departments of mainland France excluding Corsica. Data on COVID-19 tests performed and incident cases were sourced from *Santé Publique France*, the French national public health agency^22^ and for the New York State Department of Health open data repository^23^. We restricted our analysis to the time window from May 15, 2020, to November 1, 2021 (see Fig. 1a-b), to avoid the very first months of the pandemic, when testing was limited, and sharp change in incidence patterns brought about by the emergence of the Omicron variant^24,25^. Corresponding daily rainfall data were obtained from the French national weather agency Météo-France^26^ and the US National Oceanic and Atmospheric Administration^27^. Data for New York State were provided at the county level. Data for France were provided for individual weather stations, so we averaged data from stations within the same department. Weather data were retrieved from January 1, 1998 to December 30, 2022 for France and from January 1, 2016 to September 30, 2023 for New York State.

**Figure 1:**
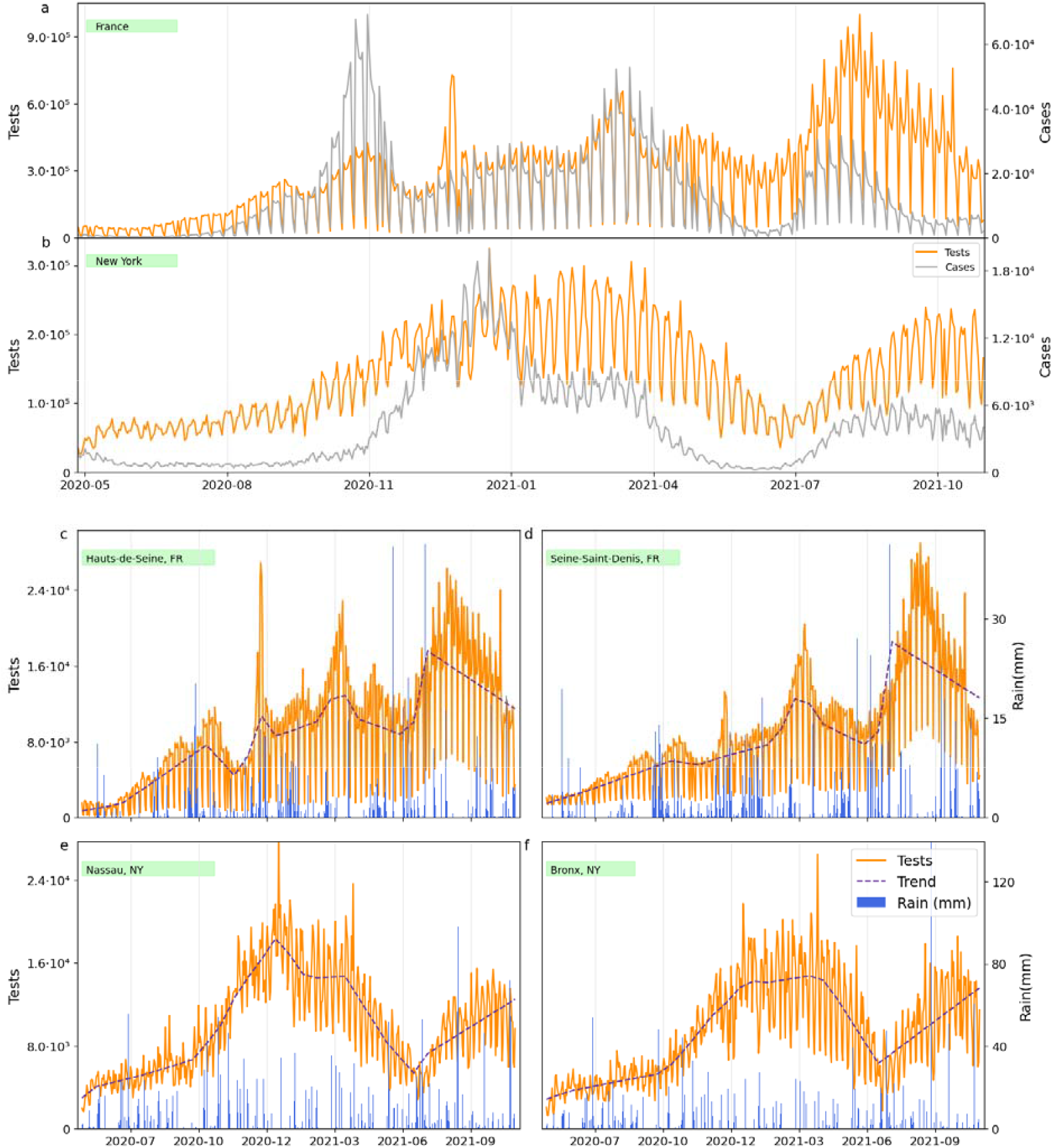
COVID-19 cases, tests, rainfall and extracted trends. (a) Daily number COVID-19 test performed (orange, left y-axis) and recorded COVID-19 cases (gray, right y-axis) in France in the study period. (b) Daily number COVID-19 test performed (orange, left y-axis) and recorded COVID-19 cases (gray, right y-axis) in New York state. (c) Daily number COVID-19 test performed (orange, left y-axis), its trend extracted from the best-performing model (see Tab. 1) and daily rainfall (blue, right y-axis) for the French department with the highest income (Hauts-de-Seine). (d) Daily number COVID-19 test performed (orange, left y-axis), its trend extracted from the best-performing model (see Tab. 1) and daily rainfall (blue, right y-axis) for the French department with the lowest income (Seine-Saint-Denis). (e) Daily number COVID-19 test performed (orange, left y-axis), its trend extracted from the best-performing model (see Tab. 2) and daily rainfall (blue, right y-axis) for the New York State county with the highest income (Nassau). (f) Daily number COVID-19 test performed (orange, left y-axis), its trend extracted from the best-performing model (see Tab. 2) and daily rainfall (blue, right y-axis) for the New York State county with the lowest income (Bronx).

Socioeconomic data were, for New York State, the county-level median household income in US dollars from the US Census Bureau (https://data.census.gov/). For France the indicator selected was the *niveau de vie* provided by the French statistical institute INSEE (https://www.insee.fr), which is defined as the disposable income in Euros of a household divided by the number of consumption units (approximately the household size). We referred to both as income indicators.

### Identification of trends and periodicities in time series

Clearly, various factors other than rainfall influenced the numbers of daily tests performed. For example, a rise in COVID-19 case counts typically triggered the increase of testing efforts^28,29^; also, logistical setups or reporting practices caused weekly periodicities in the number of tests, generally meaning that fewer tests were performed on weekends. This means that to identify the effect of rainfall we first had to adjust for exogenous trends and periodicities. To extract these, we used the forecasting model Prophet^30^. Prophet uses Markov Chain Monte Carlo to fit a time series combining (in an either additive or multiplicative way) periodic components and trends. We tuned the hyperparameters of Prophet using national (France) and state (NY) data. More details on Prophet, its use and the hyperparameter tuning are available in the Supporting Information. Then, we fit the tuned model on case data from each department (France) and county (NY), extracted time trends and weekly effects, which we then used in our main analysis. The implementation used Python version 3.12.2 and CmdStan version 2.36.0 to run Prophet. Figures 1c-f show the result of the fit with Prophet (multiplicative model) for the French departments and NY counties with the lowest and highest values of the income indicator.

### Statistical model of the effect of rainfall

We used a Poisson model to predict the daily test count as a function of rainfall. For a given area, we assumed that the expected number of tests N(t) performed on day t had the following expectation value:

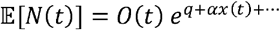

Where *O*(*t*) is the value of the trend and possibly the periodic component from the time series model (see previous section), *q* is a time-independent offset, *x*(*t*) is the daily rainfall (in mm), *α* is the coefficient to be determined, and the dots indicate other possible predictors. Then, we assumed that the likelihood of observing *N*(*t*) tests was a Poisson distribution with mean 𝔼[*N*(*t*)]. We tested four different choices for *O*(*t*): i) the trend component of the additive model; ii) the trend component of the multiplicative model; iii) the trend plus weekly component of the additive model; iv) the trend times weekly component of the multiplicative model. For cases i), ii) (i.e., without the weekly component) we also tested whether a weekly effect should be incorporated as an additional one-hot-encoded categorical variable (day of the week) in the Poisson regression.

We first fitted all these possible model choices on the national (France) and state (NY) datasets, and selected the best-performing model according to the Akaike Information Criterion. We also considered statistical models without rainfall as covariate, to check whether including rainfall was indeed meaningful in explaining the observed number of tests. The best-performing model was also fitted on each department (France) and county (NY). The models were coded and fitted in R (version 4.4.1). The coefficient *α* of the rain predictor was then translated into the more informative coefficient Δ, defined as the percentage change in daily test counts associated with additional 25 mm of rainfall: Δ= 100 · *e*^25 *α*^.

### Explaining local variations in the effect of rainfall

We analyzed spatial changes in the relationship between the effect of rainfall on testing (encoded in the value of Δ) by correlating them with two indicators: socioeconomic status and yearly rainfall profile. For the latter we chose two metrics: the average yearly number of days with more than a) 0.5 mm and b) 25 mm of rain. To account for the geographic heterogeneity in the estimate of the uncertainty of Δ we employed a weighted linear regression and estimated Pearson correlation coefficients: Point weights were proportional to the inverse of the variance of Δ as estimated by the Poisson model^31^. The weighted correlation coefficient was estimated as the slope of the linear regression after standardizing the variables.

## Results

Figure 1 provides an overview of the COVID-19 epidemic and the testing efforts in France and New York State along the study period, displaying the daily number of recorded COVID-19 cases and of performed COVID-19 tests. It also shows the daily number of tests and the daily amount of rainfall in the French departments and NY counties with the lowest and highest values of the income indicator, together with the trend component extracted with Prophet (multiplicative model).

Table 1 reports the results of the different statistical models on French national testing data. The modeling setup whereby trend and the weekly seasonality were extracted with Prophet using a multiplicative model was the one which performed best according to AIC. Importantly, rainfall indeed emerged as a meaningful predictor of the number of tests, as including it improved the lowered AIC. The estimated effect of rain (Δ) was substantially and significantly lower than zero (Δ<0%), indicating that rainfall was associated with a reduction in the number of tests performed. Specifically, we estimated an Δ=-8.7% CI=[-8.8%,-8.6%], amounting to almost a 9% reduction in the number of daily tests for every 25 mm of rainfall. The other models including rainfall, while performing worse, provided nonetheless similar estimates with reductions ranging from 8% to 11%. Table 2 reports the same analysis for New York State. The setup using the multiplicative model for Prophet, incorporating the weekly seasonality extracted and including rainfall as a predictor was again the best-performing. The effect of rain was again significantly less than zero (Δ<0), indicating that rainfall was associated with fewer tests in New York State, too. The effect was, however, lower in magnitude than in France: Δ =-4.6% CI=[-4.7%,-4.5%], an almost 5% reduction in tests for every 25 mm of rain. Worse-performing models again gave comparable results, with reductions ranging from 4.1% to 4.6%.

**Table 1:**
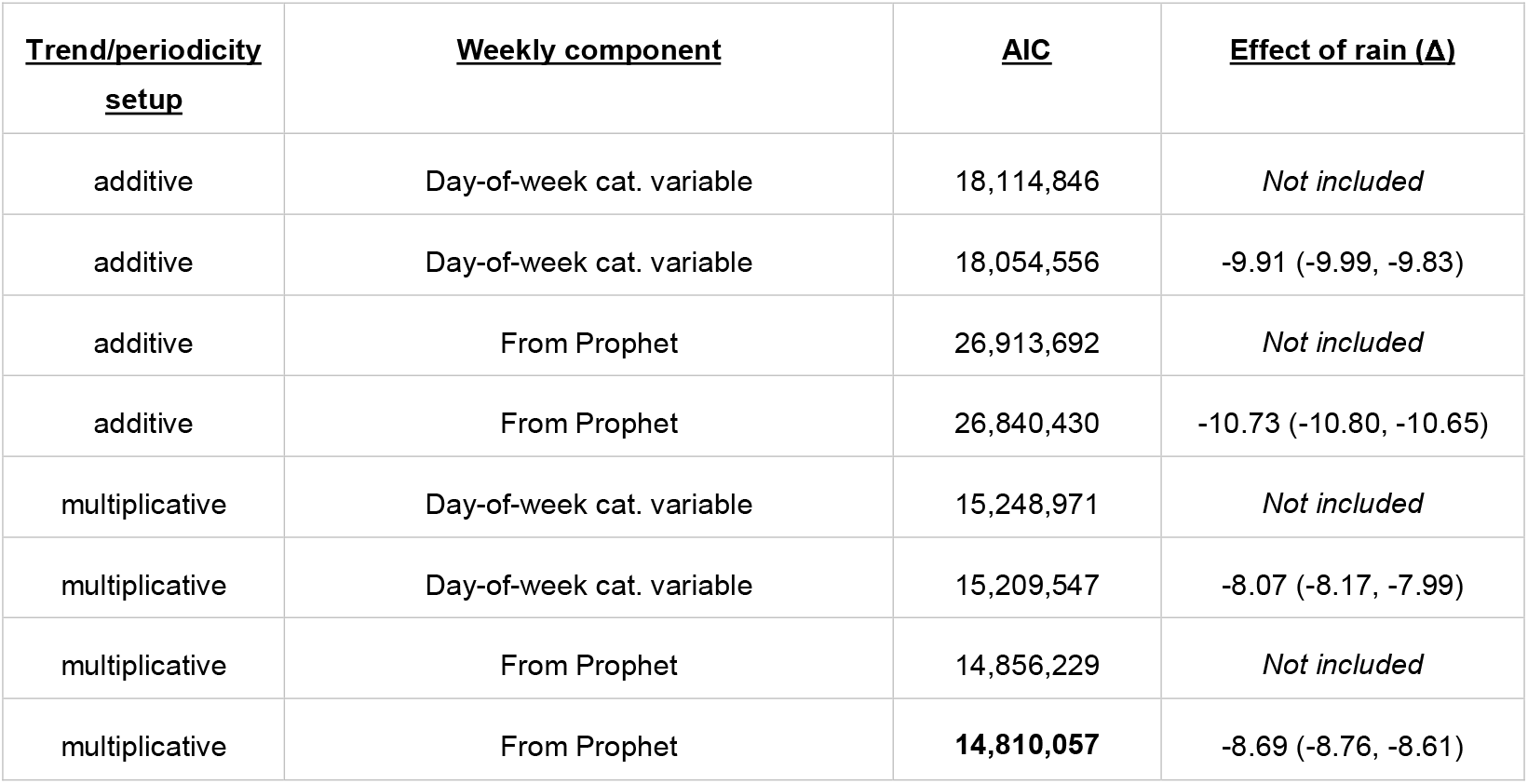
Results of the Poisson regression models for France. Each row represents a specific model setup, including the Prophet setup used (additive or multiplicative) and the way of including the weekly component. The AIC column gives the Akaike Information Criterion for each model setup. The lowest value of AIC is in bold. The Effect of rain (Δ) column reports the percentage change in the number of tests associated with 25mm of rainfall, with 95% confidence intervals in parentheses; when *not included* is reported this means that the model setup excluded rainfall as predictor, using only trend and weekly periodicity.

**Table 2:**
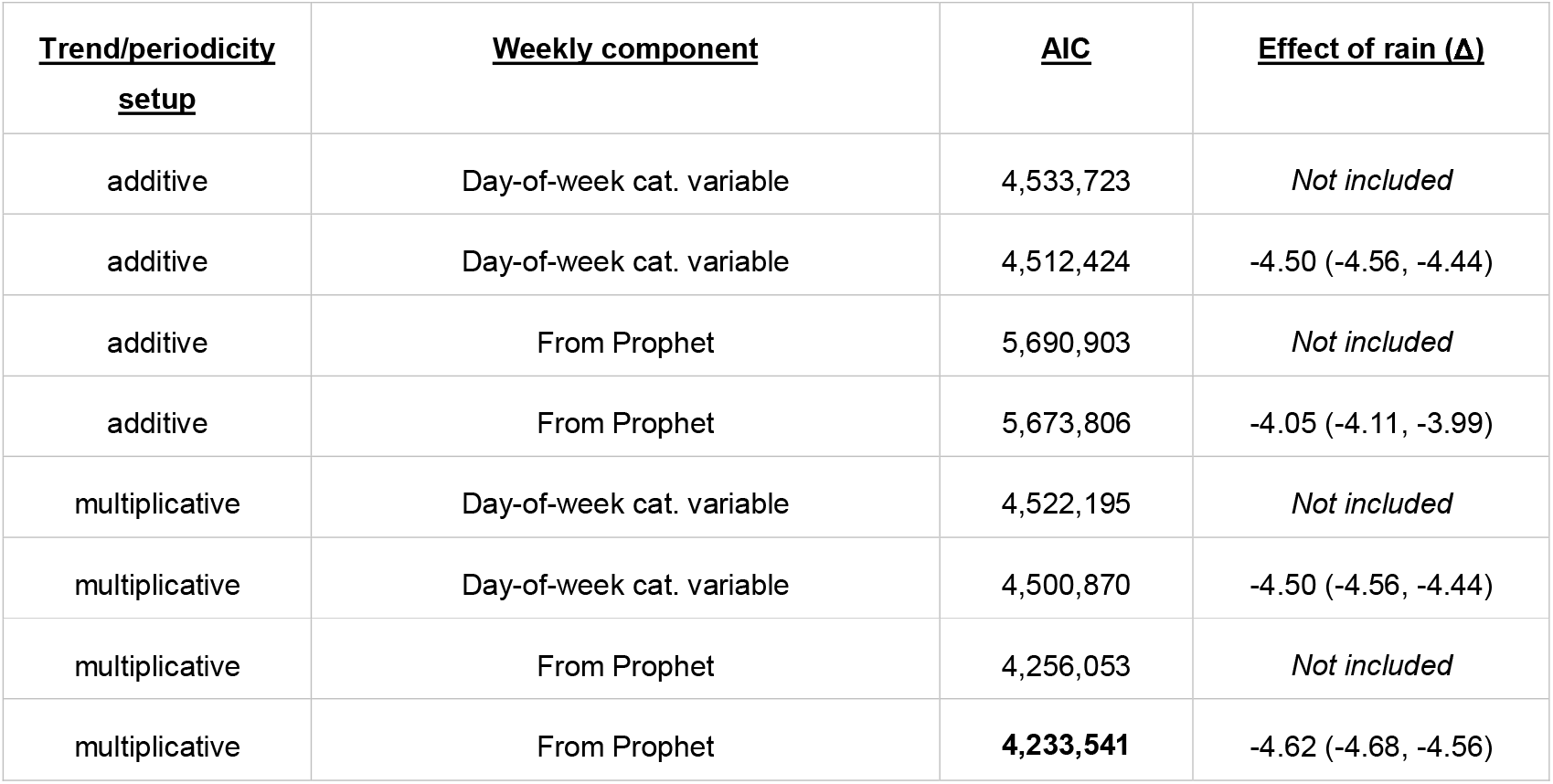
Results of the Poisson regression models for the State of New York. Each row represents a specific model setup, including the Prophet setup used (additive or multiplicative) and the way of including of the weekly component. The AIC column gives the Akaike Information Criterion for each model setup. The lowest value of AIC is in bold. The Effect of rain (Δ) column reports the percentage change in the number of tests associated with 25mm of rainfall, with 95% confidence intervals in parentheses; when *not included* is reported this means that the model setup excluded rainfall as predictor, using only trend and weekly periodicity.

| <u>Trend/periodicity setup</u> | <u>Weekly component</u> | <u>AIC</u> | <u>Effect of rain ($\Delta$)</u> |
| --- | --- | --- | --- |
| additive | Day-of-week cat. variable | 4,533,723 | *Not included* |
| additive | Day-of-week cat. variable | 4,512,424 | -4.50 (-4.56, -4.44) |
| additive | From Prophet | 5,690,903 | *Not included* |
| additive | From Prophet | 5,673,806 | -4.05 (-4.11, -3.99) |
| multiplicative | Day-of-week cat. variable | 4,522,195 | *Not included* |
| multiplicative | Day-of-week cat. variable | 4,500,870 | -4.50 (-4.56, -4.44) |
| multiplicative | From Prophet | 4,256,053 | *Not included* |
| multiplicative | From Prophet | **4,233,541** | -4.62 (-4.68, -4.56) |

The best-performing model at the national and state level was then fitted to each department and county. In France (Fig. 2a), most departments exhibited a negative relationship between rainfall and tests, compatible with the national estimate. All estimated values of Δ along with their uncertainties are reported in Supporting Information. The departments with the lowest Δ values were *Moselle* (Δ=-26.9%), *Aube* (Δ=-25.9%), *Indre* (Δ=-24.7%), *Var* (Δ=-24.5%) and *Meurthe-et-Moselle* (−22.2%). The exception to this trend was a cluster of departments in the northern part of the country, from *Ardennes* (Δ=7.2%) and *Somme* (Δ=9.0%) in the East to *Loire-Atlantique* in the West, and spanning from the coast of Normandy to the northwest of Paris (see for example *Yvelines*, Δ=12.1% and *Oise*, Δ=6.5%). In that area, additional rainfall was associated with an increase in the number of tests. It should be noted, however, that the estimate for some departments (e.g., *Somme*) had a large uncertainty. Finally, *Aisne* and *Val-d’Oise* exhibited no significant association between rainfall and tests. It should be also noted that the confidence in the estimate of Δ varied from department to department: those with high uncertainty are hatched in Fig. 2a. In New York State (Fig. 2e), as in France, most counties exhibited a negative value of Δ, compatible with the state-level fit. All estimated values of Δ along with their uncertainties are reported in Supporting Information. The strongest negative association between rainfall and testing was found in the counties of *St. Lawrence* (Δ=-17.7%), *Franklin* (Δ=-12.8%) and *Clinton* (Δ=-12.7%) in the north of the state, and *Chemung* (Δ=-13.3%) bordering Pennsylvania. The strongest positive association was found in the counties of *Rensselaer* (Δ=+4.9%) and *Columbia* (Δ=+3.7%) along the eastern border with Vermont, and *Rockland* (Δ=+4.2%), north of the Bronx. The counties with Δ>0 did not seem to cluster geographically, unlike in France. In *Cattaraugus, Chautauqua, Essex, Genesee, Greene, Jefferson, Lewis, Montgomery, Niagara, Wyoming* and *Yates* the estimate of Δ was not significantly different from zero.

**Figure 2:**
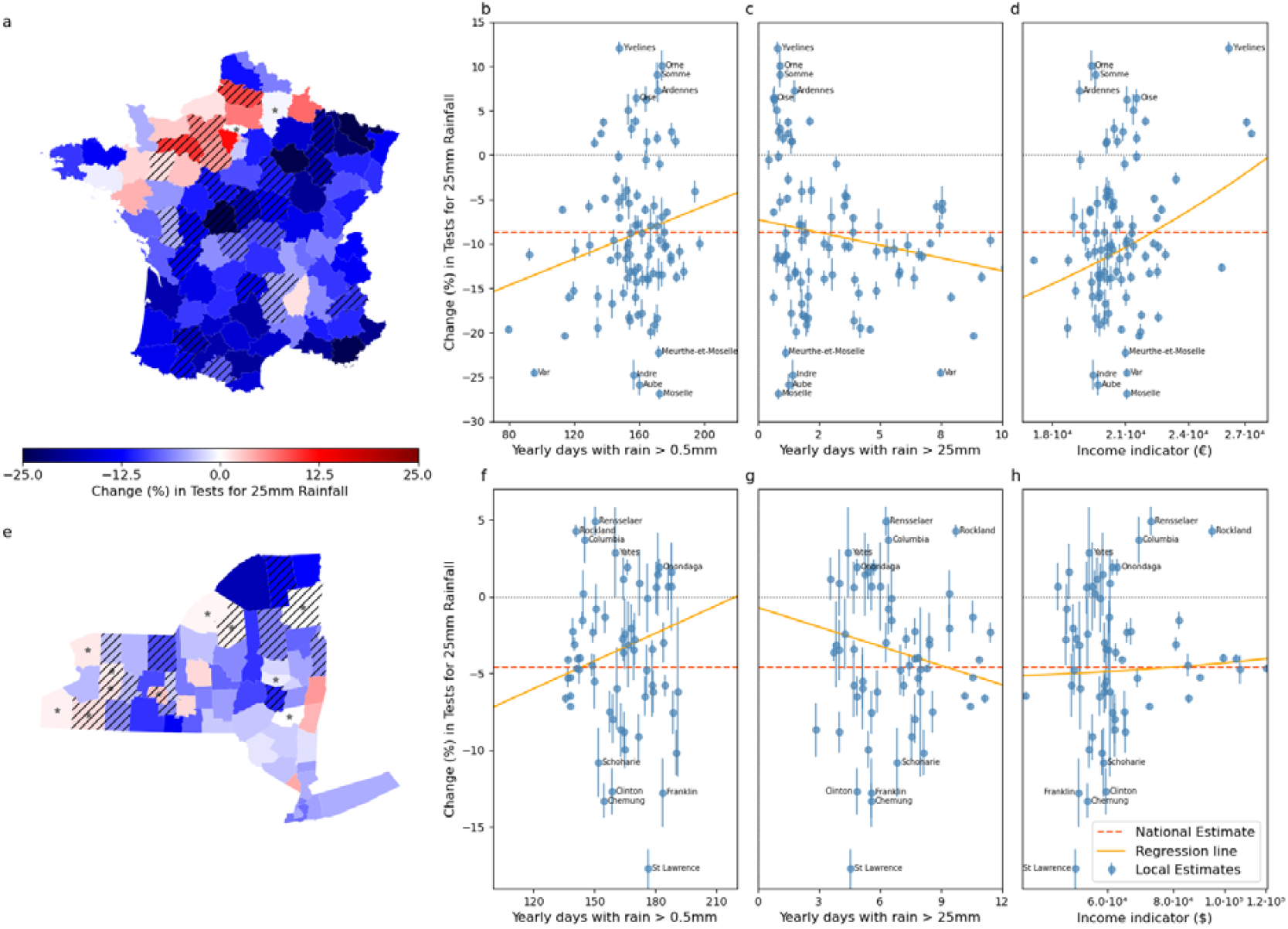
Geographical patterns of the effect of rainfall (Δ) and relationship to rainfall and socioeconomic profiles. Values of Δ come from the best-performing models (see Tab. 2). (a) Value of Δ in French departments. Hatched departments indicate that the estimated uncertainty interval on Δ is larger the third quartile of all uncertainties. Departments marked with stars indicate areas where the rainfall predictor is not statistically significantly different from zero. (b-d) scatter plots of local (French department) Δ versus the yearly number of days with more than 0.5 mm of rain (b), number of days with more than 25 mm of rain (c), and the value of the income indicator (d). (e) Value of Δ in the counties of New York State. Hatched counties indicate that the estimated uncertainty interval on Δ is larger the third quartile of all uncertainties. Counties marked with stars indicate areas where the rainfall predictor is not statistically significantly different from zero. (f-h) scatter plots of local (NY counties) Δ versus the yearly number of days with more than 0.5 mm of rain (f), number of days with more than 25 mm of rain (g), and the value of the income indicator (h). In panels (b-d) and (f-h) the red dashed line displays the national value of Δ, the orange line is the linear fit from the weighted linear regression. Numerical values of Δ displayed in (a) and (e) are reported in Supporting Information, along with their uncertainties.

We then investigated whether the local Δ was associated with the local weather profile. First, we considered the number of days in a year with more than 0.5 mm of rain. In France, the department with the largest number of such days was *Finistère* (197 days), the one with the smallest number was *Bouches-du-Rhône* (80 days). In NY, the county with the largest number of such days was *Hamilton* (191 days), the one with the smallest number was *Richmond* (136 days). We found a similar, and significantly positive, correlation between Δ and the number of those days in both France (Pearson=0.19 CI=(0.03, 0.34)) and New York State (Pearson=0.22 CI=[0.01, 0.43]) – see Fig. 2b,f. Then, we considered the number of days in a year with more than 25 mm of rain. In France, the department with the largest number of such days was *Ardèche* (13 days), the one with the smallest number was *Aisne* (<1 day). In NY, the county with the largest number of such days was *Ulster* (11 days), the one with the smallest number was *Wayne* (3 days). This time, the correlation was still similar in France and NY, but negative (Fig. 2c,g). In France, however, it was not significantly statistically different from zero (Pearson=-0.19, CI=[-0.36, 0.02]); while in New York State it was: (Pearson=-0.20, CI=[-0.35, -0.04]).

Finally, we tested the correlation between the effect of rain (Δ) and the income indicator (Fig. 2d,h). In France, a positive correlation was found: Pearson=0.26 (CI=[0.11, 0.41]). In NY, there was no statistically significant correlation: Pearson=0.05 (CI=[-0.06,0.16]).

The same analysis using the effect of rain (Δ) estimated from models other than the best-performing (see Tab. 1,2) gave similar results and is available in Supporting Information.

## Discussion

In this study, we investigated the effect of rainfall on COVID-19 testing across French departments and counties in the State of New York. We found that rainfall was significantly associated with a reduction in the number of COVID-19 tests performed, in the period from May 15, 2020, to November 1, 2021. This was true in both France and the US state of New York, but in France the effect was twice as strong. The two regions had two different COVID-19 epidemics. They had different epidemic waves (see Fig 1a) peaking at different values of incidence: 100 daily cases per 100,000 inhabitants in France, 90 in NY. They also had different public health responses, as France maintained stricter and longer-lasting restrictions on movement than New York^32–34^. The vaccination campaigns were also different, with NY starting earlier, and France catching up by June 2021. This reflected also in a different number of tests administered, peaking at around 1,300 daily tests per 100,000 inhabitants in France, 1,500 in NY. France and NY have also different weather profiles: the latter has a humid subtropical-to-continental climate with abundant precipitation throughout the year, averaging 1,000-to-1,100 mm^35,36^. Storms and tropical systems deliver frequent heavy downpours, so the state typically experiences more days with rainfall exceeding 25 mm than most parts of France. In contrast, mainland France receives about 700-to-900 mm of cumulative precipitation per year on average^37^, with a north-south gradient. These climatic differences imply that inhabitants of New York may be accustomed to more frequent and heavier rainfall, whereas many French regions - particularly in the south - experience fewer heavy rain events. These differences may help explain why the effect of rainfall on reducing the number of performed tests, albeit being present in both regions, was stronger in France: For example, heavy rainfall such as 50 mm in 24 hours was associated with almost a 20% reduction in daily tests in France, and 9% in New York State, compared to no rain. It should be noted, however, that both values, especially if compounded over multiple days, can substantially impact detection. Such disruptions directly impact public health response directly, through measures that depend on testing such as including self-isolation and quarantine, and indirectly, by impairing a correct assessment of epidemic circulation. Beyond public health, such weather-driven reduction in tests can delay individual treatment, increasing the likelihood of negative health outcomes^38^.

We found that a larger average yearly number of rainy days was associated with a smaller reduction in tests due to rainfall. This suggests that communities have adapted to keeping normal activities unperturbed when rain is common. This association was slightly stronger in New York State than in France, which is compatible with the fact this kind of adaptation is stronger in regions more accustomed to rain. We found, however, that this relationship disappeared when relating the effect on tests to the number of days of heavy rain. Here, the relationship was not significant (France) or even negative (NY), hinting that, while communities may adapt easily to moderate rainfall, they will have a hard time adapting spontaneously to extreme weather. This factor must be integrated into infectious disease preparedness in the era of extreme weather, as spontaneous community adaptation is unlikely to suffice and proactive, top-down interventions are required.

We found evidence that rainfall may reduce testing more in poorer communities, suggesting that weather-driven disruptions to epidemic monitoring are amplified in economically vulnerable populations. This aligns with studies from low- and middle-income settings showing that rainfall disproportionately disrupts healthcare access in poorer or remote communities^14,39^. But this also fits well with two more general considerations. First, the burden of COVID-19, and infectious disease epidemics in general, is unevenly distributed across socioeconomic strata^33,40^. Second, the effect of extreme weather is felt most in poorer areas^41^. The compounding of poverty, climate change, and infectious disease vulnerability highlighted by our findings underscores the urgent need for coordinated mitigation strategies to offset these intersecting risks^2^.

Our study has limitations. First, its observational design does not formally allow us to establish a causal link between rainfall and testing activity. Unknown confounders are, nonetheless, unlikely, as any such factor would need to influence rainfall itself. However, to strengthen public health guidance and inform preparedness, future work should identify the mechanisms mediating the impact of rainfall on testing, whether driven by behavioral responses to forecasts, delays in travel, or reduced transport availability. Addressing these questions will probably require individual-level studies that record personal mobility and testing decisions, moving beyond the ecological scope of our present analysis. In addition, the evidence of the association between income and the effect of rainfall is, in our study, limited to France. Our analysis does suggest this may be true in New York State, too, but the statistical power is not sufficient to make a definitive determination. Also, our type of analysis again does not allow drawing causal relationships, and our data do not allow us to control for confounders that possibly exist and need to be determined in the effort to better mitigate the compounded vulnerabilities of the poorer communities. Finally, our study is limited to two geographic settings. Despite quantitative differences and variations in statistical power, the results were, however, qualitatively consistent across two contrasting contexts. This concordance supports the potential generalizability of our findings, which warrants further investigation in other regions.

Our study provides evidence of rainfall disrupting the surveillance of respiratory epidemics by reducing diagnostic testing, especially during heavy or infrequent rain and in socioeconomically disadvantaged areas. Recognizing this vulnerability is essential to avoid delays in detection and misjudgment of epidemic trends. Integrating weather-driven risk into preparedness and implementing proactive, top-down measures can strengthen epidemic resilience under a changing climate.

## Data Availability

The studies uses previously collected and publicly available population-level data. All data repositories are cited in the manuscript.

## Acknowledgements

This study was partially supported by: Horizon Europe grant SIESTA (grant agreement no 101131957) to E.V.

## References

1. Intergovernmental Panel on Climate Change (IPCC). Climate Change 2021 – The Physical Science Basis: Working Group I Contribution to the Sixth Assessment Report of the Intergovernmental Panel on Climate Change. (Cambridge University Press, 2023).

2. Romanello, M. et al. The 2024 report of the Lancet Countdown on health and climate change: facing record-breaking threats from delayed action. The Lancet 404, 1847–1896 (2024).

3. Chen, Z., Gong, Z., Yang, S., Ma, Q. & Kan, C. Impact of extreme weather events on urban human flow: A perspective from location-based service data. Comput. Environ. Urban Syst. 83, 101520 (2020).

4. Jackson, M. L. et al. Effects of weather-related social distancing on city-scale transmission of respiratory viruses: a retrospective cohort study. BMC Infect. Dis. 21, 335 (2021).

5. Shenoy, A. et al. God is in the rain: The impact of rainfall-induced early social distancing on COVID-19 outbreaks. J. Health Econ. 81, 102575 (2022).

6. Lian, X. et al. Heat waves accelerate the spread of infectious diseases. Environ. Res. 231, 116090 (2023).

7. Arregui-García, B. et al. Disruption of outdoor activities caused by wildfire smoke shapes circulation of respiratory pathogens. PLOS Clim. 4, e0000542 (2025).

8. Lo, Y. T. E., Mitchell, D. M. & Gasparrini, A. Compound mortality impacts from extreme temperatures and the COVID-19 pandemic. Nat. Commun. 15, 4289 (2024).

9. Ascione, C. & Valdano, E. How floods may affect the spatial spread of respiratory pathogens: the case of Emilia-Romagna, Italy in May 2023. EPJ Data Sci. 14, 43 (2025).

10. Wang, Q. & Taylor, J. E. Quantifying Human Mobility Perturbation and Resilience in Hurricane Sandy. PLOS ONE 9, e112608 (2014).

11. Horanont, T., Phithakkitnukoon, S., Leong, T. W., Sekimoto, Y. & Shibasaki, R. Weather Effects on the Patterns of People’s Everyday Activities: A Study Using GPS Traces of Mobile Phone Users. PLOS ONE 8, e81153 (2013).

12. Bagrow, J. P., Wang, D. & Barabási, A.-L. Collective Response of Human Populations to Large-Scale Emergencies. PLOS ONE 6, e17680 (2011).

13. Ahmouda, A., Hochmair, H. H. & Cvetojevic, S. Using Twitter to Analyze the Effect of Hurricanes on Human Mobility Patterns. Urban Sci. 3, 87 (2019).

14. Adhvaryu, A. & Nyshadham, A. Returns to Treatment in the Formal Health Care Sector: Evidence from Tanzania. Am. Econ. J. Econ. Policy 7, 29–57 (2015).

15. Tornevi, A., Barregård, L. & Forsberg, B. Precipitation and Primary Health Care Visits for Gastrointestinal Illness in Gothenburg, Sweden. PLoS ONE 10, e0128487 (2015).

16. Susswein, Z., Rest, E. C. & Bansal, S. Disentangling the rhythms of human activity in the built environment for airborne transmission risk: An analysis of large-scale mobility data. eLife 12, e80466 (2023).

17. Jia, J. S. et al. Population flow drives spatio-temporal distribution of COVID-19 in China. Nature 582, 389–394 (2020).

18. Pullano, G. et al. Underdetection of cases of COVID-19 in France threatens epidemic control. Nature 590, 134–139 (2021).

19. Broek-Altenburg, E. van den & Atherly, A. Adherence to COVID-19 policy measures: Behavioral insights from The Netherlands and Belgium. PLOS ONE 16, e0250302 (2021).

20. Fischer, C. B. et al. Mask adherence and rate of COVID-19 across the United States. PLOS ONE 16, e0249891 (2021).

21. Petherick, A. et al. A worldwide assessment of changes in adherence to COVID-19 protective behaviours and hypothesized pandemic fatigue. Nat. Hum. Behav. 5, 1145– 1160 (2021).

22. https://www.data.gouv.fr/fr/datasets/donnees-de-laboratoires-pour-le-depistage-a-compter-du-18-05-2022-si-dep/ (Accessed August 2025).

23. https://health.data.ny.gov/Health/New-York-State-Statewide-COVID-19-Testing-Archived/xdss-u53e/about_data (Accessed August 2025).

24. Manirambona, E., Okesanya, O. J., Olaleke, N. O., Oso, T. A. & Lucero-Prisno, D. E. Evolution and implications of SARS-CoV-2 variants in the post-pandemic era. Discov. Public Health 21, 16 (2024).

25. Iuliano, A. D. Trends in Disease Severity and Health Care Utilization During the Early Omicron Variant Period Compared with Previous SARS-CoV-2 High Transmission Periods — United States, December 2020–January 2022. MMWR Morb. Mortal. Wkly. Rep. 71, (2022).

26. https://meteo.data.gouv.fr/ (Accessed August 2025).

27. https://www.ncei.noaa.gov/data/nclimgrid-daily/archive/ (Accessed August 2025).

28. Pritchard, E. et al. Detecting changes in population trends in infection surveillance using community SARS-CoV-2 prevalence as an exemplar. Am. J. Epidemiol. 193, 1848– 1860 (2024).

29. McClymont, H. et al. Internet-based Surveillance Systems and Infectious Diseases Prediction: An Updated Review of the Last 10 Years and Lessons from the COVID-19 Pandemic. J. Epidemiol. Glob. Health 14, 645–657 (2024).

30. Taylor, S. J. & Letham, B. Forecasting at scale. PeerJ Prepr. 5, e3190v2 (2017).

31. Carroll, R. J. & Ruppert, D. Transformation and Weighting in Regression. (Chapman & Hall, London, U. & New York, NY, USA, 1988).

32. Or, Z., Gandré, C., Durand Zaleski, I. & Steffen, M. France’s response to the Covid-19 pandemic: between a rock and a hard place. Health Econ. Policy Law 1–13 doi:10.1017/S1744133121000165.

33. Valdano, E., Lee, J., Bansal, S., Rubrichi, S. & Colizza, V. Highlighting socio-economic constraints on mobility reductions during COVID-19 restrictions in France can inform effective and equitable pandemic response. J. Travel Med. 28, taab045 (2021).

34. Hale, T. et al. A global panel database of pandemic policies (Oxford COVID-19 Government Response Tracker). Nat. Hum. Behav. 5, 529–538 (2021).

35. New York City Department of Environmental Protection. 2018 Drinking Water Supply and Quality Report. https://www.nyc.gov/assets/dep/downloads/pdf/water/drinking-water/drinking-water-supply-quality-report/2018-drinking-water-supply-quality-report.pdf (x2019).

36. https://www.ncei.noaa.gov/access/monitoring/climate-at-a-glance/statewide/mapping (Accessed August 2025).

37. https://donneespubliques.meteofrance.fr/?fond=produit&id_produit=117&id_rubrique=39 (Accessed August 2025).

38. Vegivinti, C. T. R. et al. Efficacy of antiviral therapies for COVID-19: a systematic review of randomized controlled trials. BMC Infect. Dis. 22, 107 (2022).

39. Stone, B., Sambo, J., Sawadogo-Lewis, T. & Roberton, T. When it rains, it pours: detecting seasonal patterns in utilization of maternal healthcare in Mozambique using routine data. BMC Health Serv. Res. 20, 950 (2020).

40. Chang, S. et al. Mobility network models of COVID-19 explain inequities and inform reopening. Nature 589, 82–87 (2020).

41. Wen, J., Song, S.-X.Cheng, Z.-Y. & Zhao, X.-X. The Impact of Extreme Weather Events on Income Inequality: Global Evidence. Emerg. Mark. Finance Trade 61, 4138– 4164 (2025).

