## Supporting Information for "Evidence of rainfall-driven disruptions in respiratory epidemics surveillance"

[1]Sorbonne Université, INSERM,
Institut Pierre Louis d'Épidémiologie et de Santé Publique,
F75012, Paris, France

* Corresponding author


[†]Current affiliation:
Dipartimento di Fisica e Astronomia "Ettore Majorana", Università di Catania, Catania, Italy.

### Trend and seasonality extraction from rainfall data

We used Prophet to estimate trend and periodic components in the rainfall time series (1). Prophet fits a time series $y(t)$ using a trend component $g(t)$, representing the non-periodic part of the time series including changepoints in overall growth, and a seasonal component $s(t)$ capturing periodic effects at different periodicities. The two components can be combined as an additive model:

$$y(t) = g(t) + s(t) + \epsilon_t$$

Or a multiplicative model:

$$y(t) = g(t) \cdot s(t) + \epsilon_t$$

Where $\epsilon_t$ is the residual error. In this study we included weekly periodicities in $s(t)$ and a piece-wise linear model for the trend.

#### *Trend*

$$g(t) = (k + \boldsymbol{a(t)}^{\boldsymbol{T}}\boldsymbol{\delta})t + (m + \boldsymbol{a(t)}^{\boldsymbol{T}}\boldsymbol{\gamma})$$

Where

- $k$: Base growth rate;
- $\boldsymbol{\delta}$: Vector of rate adjustments. For S change-points occurring at times $s_j$, $\delta_j$ represents the change in rate at $s_j$;
- $m$: Offset parameter;
- $\boldsymbol{a(t)}$**:** A binary vector indicating the presence of change-points. Its transpose is
    - $a(t)^T$ $a_j(t) = 1$ if $t \geq s_j$,
    - $a_j(t) = 0$ otherwise;
- **γ**: Adjustment parameter vector ensuring continuity of the function at change-points, calculated as: $\gamma_j = -s_j\delta_j$.

Change-points $s_j$ are automatically selected using a sparse prior on **δ**, typically $\delta_j \sim Laplace(0, \lambda)$, where λ controls the model's flexibility in adjusting its rate, fixed during hyperparameter tuning (see below).

#### *Seasonality*

$$s(t) = \sum_{n=1}^{N}\left(a_n \cos\frac{2\pi nt}{7} + b_n \sin\frac{2\pi nt}{7}\right)$$

Here, $N$ determines where the series is truncated (i.e. a low-pass filter is applied). Seasonality fitting involves estimating the $2N$ parameters $\beta = [a_1, b_1, \ldots, a_N, b_N]$. This is done by constructing a matrix of seasonality vectors for each value of $t$ in historical and future data:

$$X(t) = [cos(2\pi(1)t/7), \ldots, sin(2\pi(N)t/7)]$$

The seasonal component is then:

$$s(t) \ = \ X(t)\beta$$

In Prophet's generative model, a prior is imposed for smoothing: $\beta \sim Normal(0, \sigma^2)$.

#### *Hyperparameter tuning*

We used cross-validation to estimate the values of the hyperparameters controlling trend and seasonality encoding, namely *changepoint_prior_scale*, which controls the tendency of the model to add change points in the trends, and *seasonality_prior_scale*, which controls the smoothing of the periodic components. Cross-validation consists of an initial period used for training the model, a horizon period during which the forecast is evaluated, and a period that defines the spacing between successive cutoff dates for generating forecasts. This approach ensures a systematic evaluation of the model's forecasting performance over multiple segments of the time series. Performance is evaluated using as metrics the mean absolute error (MAE). To match the number of forecasts used in Prophet's documentation (2), we tailored the cross-validation to our tests time series, which spans 535 days. We selected an initial training period of 270 days, a forecast horizon of 44 days, and a cut-off period of 22 days (half the horizon). This configuration ensures that the cross-validation generates 12 forecasts. The same settings were applied to the tests time series for both countries at the national level. The tuning gave the following values of the hyperparameters:

- France
    - Seasonality mode: Multiplicative

        - changepoint_prior_scale: 1
        - seasonality_prior_scale: 8
    - Seasonality mode: Additive
        - changepoint_prior_scale: 0.5
        - seasonality_prior_scale: 0.5
- New York State
    - Seasonality mode: Multiplicative
        - changepoint_prior_scale: 1
        - seasonality_prior_scale: 10
    - Seasonality mode: Additive
        - changepoint_prior_scale: 1
        - seasonality_prior_scale: 5

### Supplementary results

#### *Location-specific estimated parameters*

We report the estimate of the effect of rainfall (Δ) from the best-fitting model (see Tab. 1,2 of the main text) for each French department (Tab. S1) and county in the state of New York (Tab. S2).

| Department Name | Δ (%) | 95% CI (%) |
|---|---|---|
| **Ain** | -6.1462 | [-6.69, -5.6] |
| **Aisne** | -0.51203 | [-1.59, 0.57] |
| **Allier** | -8.70949 | [-10.08, -7.32] |
| **Alpes-de-Haute-Provence** | -10.6566 | [-11.87, -9.43] |
| **Hautes-Alpes** | -10.0641 | [-11.35, -8.76] |
| **Alpes-Maritimes** | -20.3725 | [-20.76, -19.98] |
| **Ardèche** | 1.411473 | [0.81, 2.01] |
| **Ardennes** | 7.238194 | [6, 8.48] |
| **Ariège** | -7.91404 | [-9.75, -6.05] |
| **Aube** | -25.867 | [-27.03, -24.69] |
| **Aude** | -19.4383 | [-20.62, -18.25] |
| **Aveyron** | -18.6268 | [-19.68, -17.56] |
| **Bouches-du-Rhône** | -19.6363 | [-20, -19.27] |
| **Calvados** | 2.660675 | [1.62, 3.71] |
| **Cantal** | -13.8187 | [-15.08, -12.54] |
| **Charente** | -13.0509 | [-14.48, -11.6] |
| **Charente-Maritime** | -7.86166 | [-8.96, -6.75] |
| **Cher** | -17.8147 | [-19.35, -16.26] |
| **Corrèze** | -10.526 | [-11.54, -9.51] |
| **Côte-d'Or** | -19.8838 | [-20.75, -19.02] |
| **Côtes-d'Armor** | -13.0654 | [-14.08, -12.04] |
| **Creuse** | -6.96482 | [-9.14, -4.74] |
| **Dordogne** | -14.2766 | [-15.45, -13.09] |
| **Doubs** | -11.3524 | [-11.97, -10.73] |

| | | |
|---|---|---|
| **Drôme** | -5.77494 | [-6.51, -5.03] |
| **Eure** | 6.326435 | [4.95, 7.72] |
| **Eure-et-Loir** | 5.133653 | [3.39, 6.9] |
| **Finistère** | -9.93606 | [-10.73, -9.14] |
| **Gard** | -6.15668 | [-6.57, -5.74] |
| **Haute-Garonne** | -18.2299 | [-18.83, -17.63] |
| **Gers** | -16.6818 | [-18.29, -15.05] |
| **Gironde** | -18.0235 | [-18.52, -17.52] |
| **Hérault** | -15.9598 | [-16.51, -15.4] |
| **Ille-et-Vilaine** | 1.964683 | [1.2, 2.73] |
| **Indre** | -24.7263 | [-26.35, -23.08] |
| **Indre-et-Loire** | -11.2676 | [-12.46, -10.07] |
| **Isère** | -9.90681 | [-10.34, -9.47] |
| **Jura** | -9.57466 | [-10.35, -8.79] |
| **Landes** | -14.0284 | [-14.84, -13.21] |
| **Loir-et-Cher** | -11.642 | [-13.25, -10.01] |
| **Loire** | -4.50142 | [-5.14, -3.86] |
| **Haute-Loire** | -4.64044 | [-6.03, -3.24] |
| **Loire-Atlantique** | 3.807076 | [3.27, 4.35] |
| **Loiret** | -8.75444 | [-9.79, -7.71] |
| **Lot** | -10.411 | [-11.66, -9.15] |
| **Lot-et-Garonne** | -9.57741 | [-10.75, -8.39] |
| **Lozère** | -5.38824 | [-7.23, -3.53] |
| **Maine-et-Loire** | -5.24663 | [-6.31, -4.18] |
| **Manche** | -4.06339 | [-5.25, -2.86] |
| **Marne** | -16.0148 | [-16.96, -15.06] |
| **Haute-Marne** | -18.383 | [-19.93, -16.82] |
| **Mayenne** | 1.607431 | [0.16, 3.06] |
| **Meurthe-et-Moselle** | -22.197 | [-22.87, -21.52] |
| **Meuse** | -19.0559 | [-20.41, -17.69] |
| **Morbihan** | -0.96596 | [-1.81, -0.12] |

| | | |
|---|---|---|
| **Moselle** | -26.8828 | [-27.46, -26.3] |
| **Nièvre** | -13.4829 | [-15.27, -11.67] |
| **Nord** | -6.43004 | [-6.94, -5.91] |
| **Oise** | 6.476802 | [5.48, 7.49] |
| **Orne** | 10.08101 | [8.38, 11.8] |
| **Pas-de-Calais** | -11.8577 | [-12.53, -11.18] |
| **Puy-de-Dôme** | -10.1797 | [-10.91, -9.44] |
| **Pyrénées-Atlantiques** | -13.7635 | [-14.39, -13.13] |
| **Hautes-Pyrénées** | -13.4405 | [-14.84, -12.02] |
| **Pyrénées-Orientales** | -15.2707 | [-16.27, -14.26] |
| **Bas-Rhin** | -13.2496 | [-13.82, -12.68] |
| **Haut-Rhin** | -10.2634 | [-10.88, -9.64] |
| **Rhône** | -7.0328 | [-7.38, -6.68] |
| **Haute-Saône** | -8.6375 | [-9.82, -7.44] |
| **Saône-et-Loire** | -13.8715 | [-14.75, -12.98] |
| **Sarthe** | 3.002194 | [1.9, 4.12] |
| **Savoie** | -13.07 | [-13.94, -12.19] |
| **Haute-Savoie** | -12.6828 | [-13.14, -12.22] |
| **Paris** | 2.502537 | [2.14, 2.87] |
| **Seine-Maritime** | 1.530849 | [0.74, 2.33] |
| **Seine-et-Marne** | -11.1747 | [-11.79, -10.55] |
| **Yvelines** | 12.09169 | [11.35, 12.84] |
| **Deux-Sèvres** | -3.97273 | [-5.44, -2.49] |
| **Somme** | 9.054456 | [7.69, 10.44] |
| **Tarn** | -15.4924 | [-16.44, -14.53] |
| **Tarn-et-Garonne** | -15.8783 | [-17.12, -14.62] |
| **Var** | -24.5145 | [-24.96, -24.07] |
| **Vaucluse** | -11.1849 | [-11.94, -10.43] |
| **Vendée** | -7.77478 | [-8.73, -6.81] |
| **Vienne** | -13.8034 | [-15.16, -12.43] |
| **Haute-Vienne** | -5.64273 | [-6.7, -4.58] |

| | | |
|---|---|---|
| **Vosges** | -10.8599 | [-11.71, -10.01] |
| **Yonne** | -11.2629 | [-12.71, -9.79] |
| **Territoire de Belfort** | -7.9546 | [-9.17, -6.73] |
| **Essonne** | -2.63654 | [-3.34, -1.93] |
| **Hauts-de-Seine** | 3.725502 | [3.25, 4.2] |
| **Seine-Saint-Denis** | -11.8111 | [-12.28, -11.34] |
| **Val-de-Marne** | -4.89253 | [-5.41, -4.37] |
| **Val-d'Oise** | -0.12645 | [-0.79, 0.54] |

**Table S1: Estimate of the effect of rainfall in French departments from the best-fitting model.**

| County Name | Δ (%) | 95% CI |
|---|---|---|
| **Albany** | -5.3086 | [-5.94, -4.67] |
| **Allegany** | -5.77949 | [-7.83, -3.7] |
| **Bronx** | -6.43835 | [-6.64, -6.23] |
| **Broome** | -2.71989 | [-3.43, -2.01] |
| **Cattaraugus** | 1.61815 | [-0.16, 3.42] |
| **Cayuga** | -6.50175 | [-7.77, -5.22] |
| **Chautauqua** | 0.642014 | [-0.9, 2.2] |
| **Chemung** | -13.307 | [-14.43, -12.17] |
| **Chenango** | -4.79342 | [-6.41, -3.16] |
| **Clinton** | -12.6796 | [-14.19, -11.15] |
| **Columbia** | 3.666944 | [2.17, 5.17] |
| **Cortland** | -6.19387 | [-7.58, -4.8] |
| **Delaware** | -2.78705 | [-4.49, -1.07] |
| **Dutchess** | -1.55544 | [-2.1, -1.01] |
| **Erie** | -3.40455 | [-3.95, -2.86] |
| **Essex** | -0.10914 | [-2.33, 2.14] |
| **Franklin** | -12.767 | [-14.95, -10.54] |
| **Fulton** | -2.0576 | [-3.6, -0.5] |
| **Genesee** | 0.88889 | [-1.26, 3.07] |

| | | |
|---|---|---|
| **Greene** | 0.169667 | [-1.37, 1.72] |
| **Hamilton** | -6.21965 | [-11.62, -0.62] |
| **Herkimer** | -10.1513 | [-11.61, -8.68] |
| **Jefferson** | 0.580003 | [-1.1, 2.28] |
| **Kings** | -4.08865 | [-4.23, -3.95] |
| **Lewis** | 0.667675 | [-2.18, 3.58] |
| **Livingston** | -3.49721 | [-5.43, -1.54] |
| **Madison** | -7.55811 | [-8.95, -6.16] |
| **Monroe** | -3.64158 | [-4.2, -3.08] |
| **Montgomery** | -0.82296 | [-2.47, 0.84] |
| **Nassau** | -4.6751 | [-4.92, -4.43] |
| **New York** | -5.23939 | [-5.39, -5.09] |
| **Niagara** | 1.135778 | [-0.25, 2.54] |
| **Oneida** | -5.79342 | [-6.5, -5.09] |
| **Onondaga** | 1.903683 | [1.36, 2.45] |
| **Ontario** | -8.83928 | [-10.16, -7.51] |
| **Orange** | -3.08716 | [-3.54, -2.63] |
| **Orleans** | -3.14321 | [-5.93, -0.3] |
| **Oswego** | -3.02014 | [-4.31, -1.72] |
| **Otsego** | -9.1487 | [-10.65, -7.63] |
| **Putnam** | -4.74035 | [-5.65, -3.83] |
| **Queens** | -7.15319 | [-7.32, -6.98] |
| **Rensselaer** | 4.883071 | [3.96, 5.82] |
| **Richmond** | -6.62386 | [-6.91, -6.34] |
| **Rockland** | 4.245954 | [3.82, 4.67] |
| **St. Lawrence** | -17.7317 | [-18.99, -16.46] |
| **Saratoga** | -4.48435 | [-5.21, -3.76] |
| **Schenectady** | -2.28602 | [-3.16, -1.41] |
| **Schoharie** | -10.8064 | [-13.03, -8.55] |
| **Schuyler** | -5.98464 | [-8.62, -3.3] |
| **Seneca** | -2.44446 | [-4.73, -0.13] |

| | | |
|---|---|---|
| **Steuben** | -9.98092 | [-11.19, -8.76] |
| **Suffolk** | -4.03009 | [-4.27, -3.79] |
| **Sullivan** | -1.32656 | [-2.37, -0.28] |
| **Tioga** | -7.95749 | [-9.55, -6.35] |
| **Tompkins** | 1.885577 | [1.41, 2.36] |
| **Ulster** | -2.34961 | [-2.98, -1.72] |
| **Warren** | -7.51672 | [-8.87, -6.16] |
| **Washington** | -5.53756 | [-7.27, -3.79] |
| **Wayne** | -8.68254 | [-10.45, -6.89] |
| **Westchester** | -4.00162 | [-4.27, -3.74] |
| **Wyoming** | 1.422471 | [-1.27, 4.17] |
| **Yates** | 2.84998 | [-0.08, 5.82] |

**Table S2: Estimate of the effect of rainfall in NY State counties from the best-fitting model.**

#### *Alternative statistical models*

According to Tab. 1,2 of the main paper, the best-performing model for both countries is the one that includes the weekly component from Prophet as an offset and uses precipitation as a predictor in the Poisson regression. In Figures S1, S2 we report the association between rainy days and socioeconomic status v the effect of rainfall, for the other statistical models in Tab. 1,2 which included rainfall.

For France (Fig. S1), the geographical pattern of the effect of rainfall (Δ) remains evident in the model using as offset the trend obtained from the multiplicative seasonality model of Prophet, with the days of the week included as a one-hot-encoded categorical variable. In this model, Δ continues to show a weak negative correlation with the annual number of days with rainfall above 25 mm (correlation = -0.170, 95% confidence interval (CI): -0.337 to -0.004), and a positive correlation with the income indicator (correlation = 0.252, 95% CI: 0.111 to 0.394). However, the correlation with the number of days with rainfall exceeding 0.5mm is no longer statistically significant. Notably, the three departments with the highest and lowest Δ values in this model also appear among the top five highest and lowest values in the best-performing model.

In the additive models (i.e., models with the trend extracted from the additive version of Prophet), the only statistically significant relationship observed for Δ is with the income indicator. Specifically, the Pearson correlation is 0.339 (95% CI: 0.201 to 0.476) when using the Prophet's weekly component as an offset, and 0.319 (95% CI: 0.177 to 0.460) when using the day of the week as a predictor.

In the state of New York (Fig. S2), no obvious geographical pattern is visible as in the best-fitting model; however, some regions exhibit consistent trends across all models. Notably, the southeastern peninsula consistently shows a weak blue shade, while the counties of *Columbia, Rensselaer, Rockland*, and *Onondaga* display a persistent red shade across all trained models, indicating a level of consistency in the results.

None of the models reveal a significant relationship between the percentage change of tests associated with 25 mm of rain (Δ) and the income indicator, reinforcing the findings of the best-performing model. Conversely, the Δ of all the models show a moderate but significant negative correlation with the annual number of days registering more than 25 mm of precipitation. The percentage change of tests (Δ) obtained from the multiplicative model, where the day of the week is used as predictor, has a Pearson correlation of -0.221 with a 95% confidence interval of (-0.378, -0.065) with the yearly number of days with rain exceeding 25 mm. For the additive model, which includes the Prophet's weekly component as offset, Δ shows a correlation of -0.163 (95% CI: -0.320, -0.006), while for the additive model with the day of the week as predictor, Δ presents a correlation of -0.200 (95% CI: -0.358, -0.043). Finally, the correlation with the annual number of days experiencing more than 0.5mm of precipitation is significant for the multiplicative models. The Pearson correlation of Δ for the model using the day of the week as predictor is 0.271 (95% CI: 0.064, 0.479. Among the additive models, significance is observed only in the model that includes weekly seasonality as predictor, with a Pearson correlation of 0.230 (95% CI: 0.021, 0.439). Regarding the counties with the highest and lowest values of Δ, they consistently rank among the five highest and lowest values in the best-performing model, further demonstrating the robustness of the results.

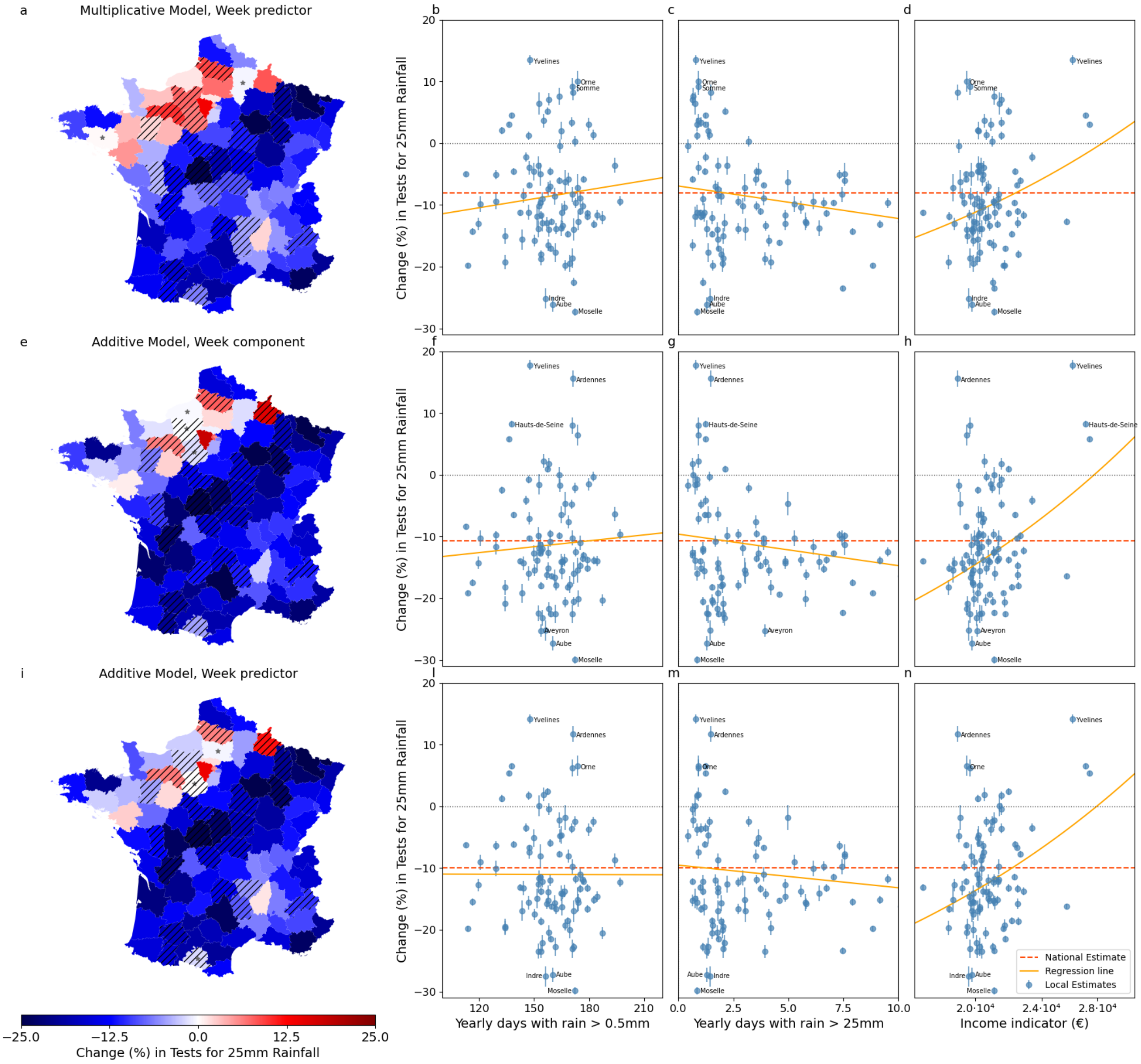


**Figure S1: Geographical patterns of the effect of rainfall (Δ) and relationship to rainfall and socioeconomic profiles in France – alternative models.** Each row of the figure is equivalent to panels (a-d) of Fig. 2 in the main text, but with values of Δ from a model other than the best-fitting one (see Tab. 1 of the main text). The model is indicated on top of the map. (a,e,i) Value of Δ in French departments. Hatched departments indicate that the estimated uncertainty interval on Δ is larger than the third quartile of all uncertainties. Departments marked with stars indicate areas where the rainfall predictor is not statistically significantly different from zero. (b-d, f-h, l-n) scatter plots of local Δ versus the yearly number of days with more than 0.5 mm of rain (b,f,l), number of days with more than 25 mm of rain (c,g,m), and the value of the income indicator (d,h,n). The red dashed line displays the national value of Δ, the orange line is the linear fit from the weighted linear regression.

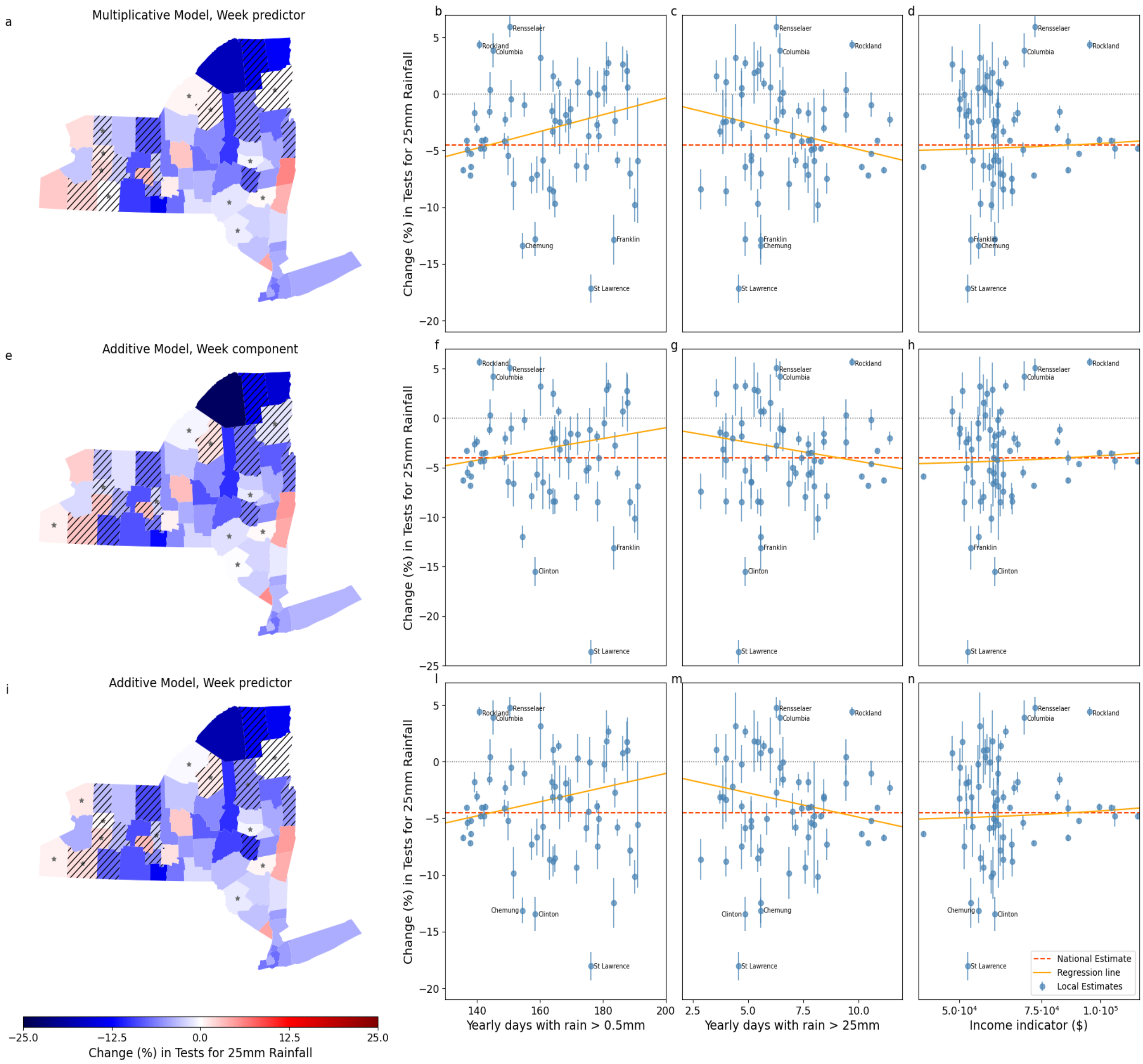


**Figure S2: Geographical patterns of the effect of rainfall (Δ) and relationship to rainfall and socioeconomic profiles in NY State – alternative models.** Each row of the figure is equivalent to panels (e-h) of Fig. 2 in the main text, but with values of Δ from a model other than the best-fitting one (see Tab. 2 of the main text). The model is indicated on top of the map. (a,e,i) Value of Δ in NY counties. Hatched counties indicate that the estimated uncertainty interval on Δ is larger the third quartile of all uncertainties. Counties marked with stars indicate areas where the rainfall predictor is not statistically significantly different from zero. (b-d, f-h, l-n) scatter plots of local Δ versus the yearly number of days with more than 0.5 mm of rain (b,f,l), number of days with more than 25 mm of rain (c,g,m), and the value of the income indicator (d,h,n). The red dashed line displays the national value of Δ, the orange line is the linear fit from the weighted linear regression.

#### References

1. Taylor SJ, Letham B. Forecasting at scale. PeerJ Preprints; 2017 Sep . Report No.: e3190v2. Available from: https://peerj.com/preprints/3190

2. Prophet. 2024. Diagnostics. Available from: http://facebook.github.io/prophet/docs/diagnostics.html
